# Serial interval and incubation period estimates of monkeypox virus infection in 12 U.S. jurisdictions, May – August 2022

**DOI:** 10.1101/2022.10.26.22281516

**Authors:** Zachary J. Madewell, Kelly Charniga, Nina B. Masters, Jason Asher, Lily Fahrenwald, William Still, Judy Chen, Naama Kipperman, David Bui, Meghan Shea, Lori Saathoff-Huber, Shannon Johnson, Khalil Harbi, Abby L. Berns, Taidy Perez, Emily Gateley, Ian H. Spicknall, Yoshinori Nakazawa, Thomas L. Gift, 2022 Monkeypox Outbreak Response Team

## Abstract

Using data collected by 12 U.S. health departments, we report mean estimated serial interval for monkeypox virus infection of 8.5 (95% CrI: 7.3 – 9.9) days for symptom onset from 57 case pairs and mean estimated incubation period of 5.6 (4.3 – 7.8) days from 35 case pairs for symptom onset.

**One sentence summary:** We report the mean estimated serial interval for monkeypox virus infection of 8.5 (95% CrI: 7.3 – 9.9) days for symptom onset from 57 case pairs and mean estimated incubation period of 5.6 (4.3 – 7.8) days from 35 case pairs for symptom onset.

---

Since May 6, 2022, monkeypox cases have been reported across the globe. According to the U.S. Centers for Disease Control and Prevention (CDC), there have been 75,568 confirmed monkeypox cases and 34 deaths in 109 locations across both historically endemic and non-endemic regions through October 25, 2022 (1). Monkeypox symptoms usually start within three weeks of exposure to monkeypox virus (MPXV) and may include fever, headache, chills, swollen lymph nodes, and exhaustion (1). A rash usually develops within one to four days after onset of symptoms. MPXV is transmitted via close contact with infectious rash, scabs, or body fluids, respiratory droplets during prolonged face-to-face contact, and fomites such as clothing, towels, or bedding (1). Transmission in the current outbreak has occurred primarily through close physical contact associated with sexual activities among gay, bisexual, and other men who have sex with men. Transmission of MPXV is possible from symptom onset until all scabs have fallen off and fully healed (2).

The serial interval is defined as the time between symptom onset in a primary case and symptom onset in the secondary case and depends on the incubation period (the time from a person’s infection to the onset of signs and symptoms) (3), the degree of infectiousness of an index case, and population contact patterns. The serial interval is critical for estimating the effective reproduction number, *R*_*t*_, and forecasting incidence, which are both important for understanding the course of an outbreak and impact of interventions (e.g., antivirals and vaccines). In the current outbreak, many patients report multiple anonymous sex partners or attendance at large events, such as festivals, in the three weeks prior to symptom onset which has complicated efforts to identify primary and secondary case pairs. Using preliminary data from 17 monkeypox case pairs in the UK, authors estimated the mean serial interval to be 9.8 days with high uncertainty (95% credible interval [CrI]: 5.9 – 21.4) (4). An investigation of 16 primary/secondary case pairs in Italy estimated the mean generation time, or time between infection of primary and secondary cases, to be 12.5 days (95% CrI: 7.5 – 17.3) (5). Although the serial interval and the generation time theoretically have the same mean, they may not have the same variance (6). Here, we estimate the serial interval and incubation period for both symptom onset and rash onset for MPXV infection for the current outbreak in the United States.

## The Study

Data on symptom and rash onset dates for primary and secondary case pairs as well as the type of contact that occurred between pairs (i.e., sexual or intimate contact; face-to-face contact; caregiving; shared bedding; unknown) were compiled by 12 state and local health departments (listed in acknowledgments). Cases were only included if there was a high degree of certainty that the secondary case was infected by the primary case, meeting the four criteria below. Patient interviews conducted during contact tracing confirmed that secondary cases did not: (1) report exposure to multiple potential primary cases; (2) have more than one sexual partner in the three weeks prior to symptom onset; (3) have symptom onset on the same day as the primary case; or (4) attend a festival, bath house, sex party, or other crowded event in the three weeks prior to symptom onset and had minimal or no clothing during the event.

Days between onset of any monkeypox symptoms and days between rash onset in the primary and secondary cases were calculated for each case pair. The EpiEstim package (version 4.1.2) in R software (R Foundation for Statistical Computing) was used to estimate the distribution of the serial interval for known primary and secondary case pairs using Bayesian methods for both symptom and rash onset (7). Log-normal, Weibull, and gamma distributions were fitted to the difference-in-days data. Akaike’s information criterion (AIC) was used to compare fits, and the model with the lowest AIC value was selected. The serial interval was estimated for symptom onset and rash onset using 50,000 Markov Chain Monte Carlo (MCMC) samples and a burn-in of 10,000 samples. Convergence of MCMC samples was assessed by the Gelman–Rubin statistic. We did not adjust for right-truncation of the data as cases from four months of the outbreak were included (Figure S1). Fifty-seven case pairs met the inclusion criteria (Figure 1). Dates of symptom onset among primary cases ranged from May 11 to August 13, 2022. Forty pairs out of 57 had rash onset dates for primary and secondary cases. Type of contact for the case pairs can be found in Table S1. The gamma distribution provided the best fit to the serial interval data. The overall mean estimated serial interval for symptom onset was 8.5 (95% CrI: 7.3 – 9.9) days (SD: 5.0 [95% CrI: 4.0 – 6.4] days) and for rash onset was 7.0 (95% CrI: 5.8 – 8.4) days (SD: 4.2 [95% CrI: 3.2 – 5.6] days) (Table 1, Table S2).

**Figure 1.**
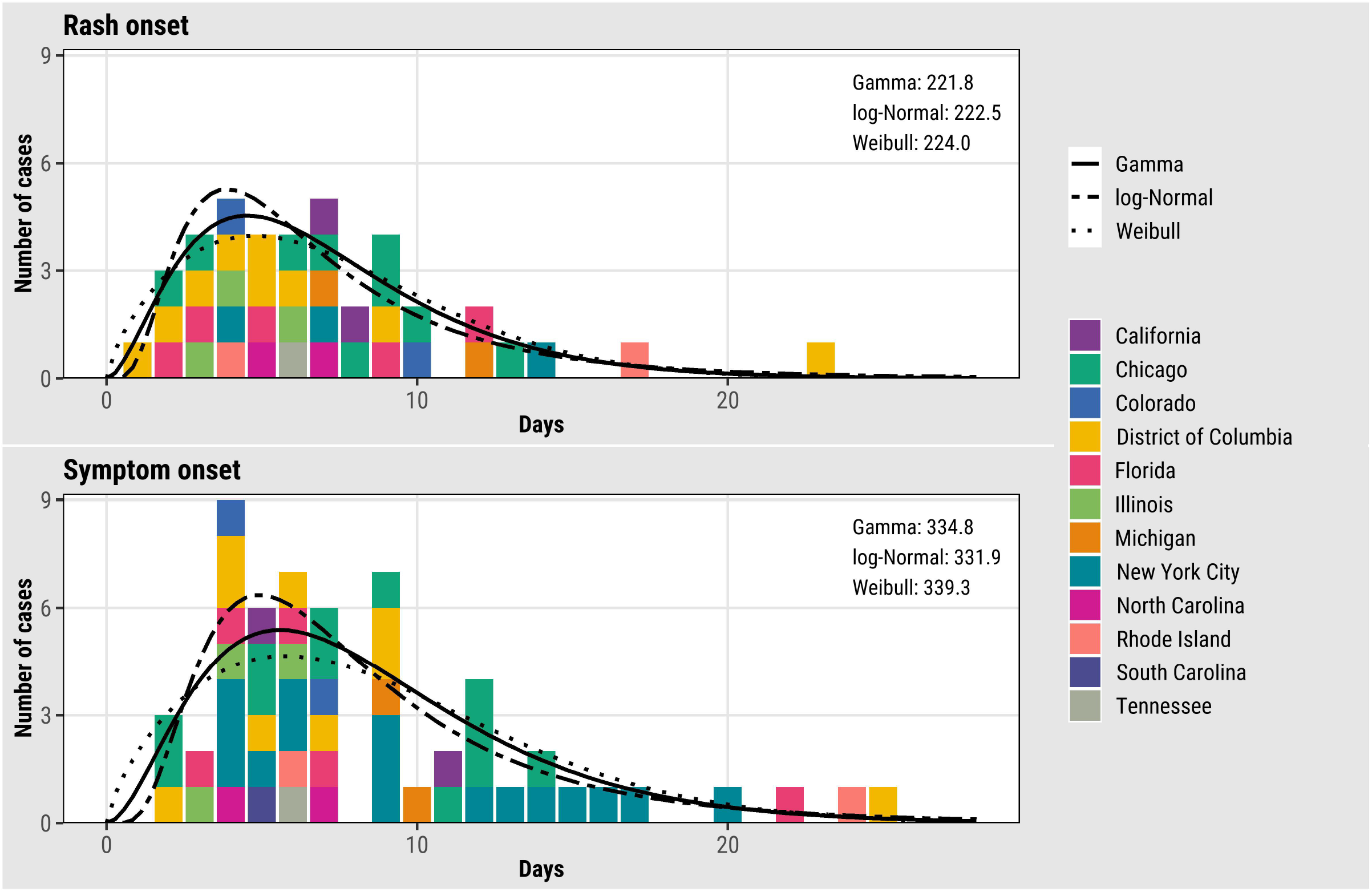
Empirical and fitted distributions of the serial intervals for rash (N=40) and symptom (N=57) onset for monkeypox virus, United States, May – August 2022. Colors represent U.S. jurisdictions (N = 12). AIC values for each model are shown inside the plots in the upper right-hand corner.

**Figure 2.**
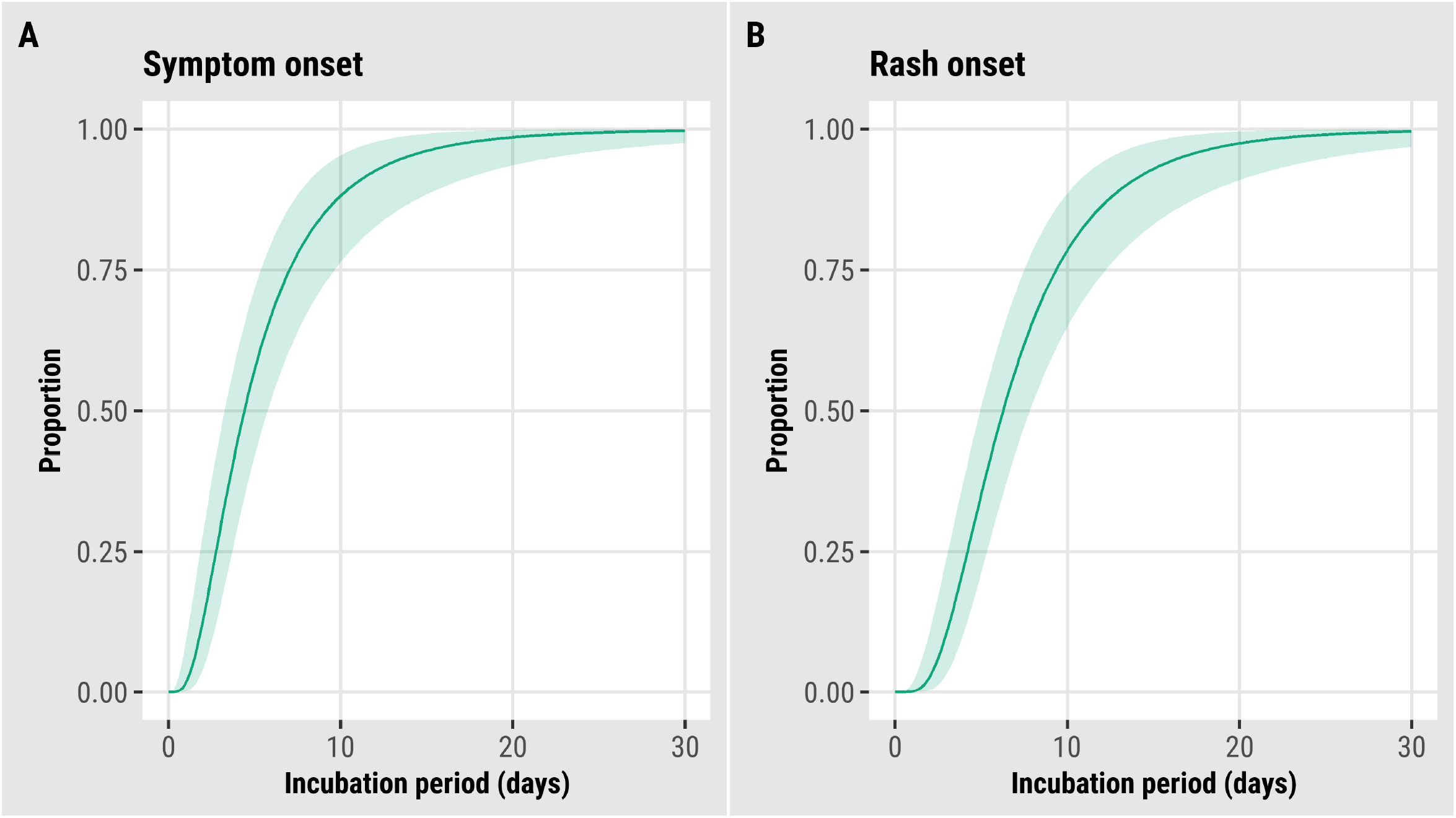
Updated estimated cumulative density functions according to a log-normal distribution of monkeypox virus incubation periods, United States, May – August 2022. (A) symptom onset (N = 36) and (B) rash onset (N = 35).

**Table 1.** Estimated incubation period and serial interval of monkeypox virus, United States, May – August 2022

|  | Incubation period in days |  |  | Serial interval in days |  |  |
| --- | --- | --- | --- | --- | --- | --- |
|  | Mean (95% CrI) | Standard deviation (95% CrI) | Sample size (cases) | Mean (95% CrI) | Standard deviation (95% CrI) | Sample size (case pairs) |
| Symptom onset | 5.6 (4.3 – 7.8) | 4.4 (2.8 – 8.7) | 36 | 8.5 (7.3 – 9.9) | 5.0 (4.0 – 6.4) | 57 |
| Rash onset | 7.5 (6.0 – 9.8) | 4.9 (3.2 – 8.8) | 35 | 7.0 (5.8 – 8.4) | 4.2 (3.2 – 5.6) | 40 |

We added nine cases from the District of Columbia and five cases from Florida to the MPXV incubation period analysis first reported by Charniga et al. (only U.S. cases were included, unlike in the preprint) (8). Of the new cases, 10 were exposed during a single day. The mean incubation period from exposure to symptom onset for 36 cases was 5.6 (95% CrI: 4.3 – 7.8) days (SD: 4.4 [95% CrI: 2.8 – 8.7] days), while the mean incubation period from exposure to rash onset for 35 cases was 7.5 (95% CrI: 6.0 – 9.8) days (SD: 4.9 [95% CrI: 3.2 – 8.8] days). We found a statistically significant correlation between the serial intervals and incubation periods in secondary cases that were included in both analyses (N = 15, Figure S2).

## Conclusions

Determining the serial interval of a pathogen can inform our understanding of the timing of transmission relative to symptom onset. The serial interval of 8.5 days for symptom onset was similar to an estimate from the UK of 9.8 days after correcting for right-truncation (4), but shorter than the generation time of 12.5 days reported from Italy (5). We also found a serial interval of 7.0 days for rash onset. We are unaware of other studies reporting rash serial interval for monkeypox. The estimated serial interval for symptom onset was longer than the incubation period (5.6 days), suggesting most transmission occurred after the onset of symptoms in the primary case. Conversely, the serial interval for rash onset (7.0 days) was slightly shorter than the rash incubation period (7.5 days), which may suggest some pre-rash transmission; (indeed, there were instances in the observed data where secondary cases were exposed before onset of reported rash in the primary case); however, the credible intervals for the estimates overlap. The serial interval for symptom onset ranged from two to 25 days. This wide range may be attributed in part to variability in the nature and intensity of contact.

This study had several limitations. First, precise ascertainment of symptom and rash onset dates is critical for serial interval estimation, but initial monkeypox symptoms are often non-specific and may be unrelated to MPXV infection. Second, despite careful selection of linked primary/secondary case pairs, exposure from additional unknown sources may have occurred. Third, there may have been social desirability bias in the self-reported exposures prior to infection. Fourth, serial interval may vary by age, comorbidity status, vaccination status, or contact type (route of exposure); we did not stratify our analysis by these factors due to limited data. Fifth, we excluded secondary cases that had symptom onset on the same day as or before the primary case to ensure a high degree of confidence linking case pairs; however, the serial interval could be negative (9). Sixth, the serial interval for rash onset could be biased if rash is more quickly identified in the secondary case because of case finding/investigation of the primary case.

Our estimate of the serial interval includes more case pairs than have been reported previously, and we include estimates for rash onset, which may be more reliable than initial symptoms. These early data from the monkeypox outbreak in the United States demonstrate the mean serial interval is 8.5 days for symptom onset.

## Data Availability

All data produced in the present work are contained in the manuscript.

## Biographical sketch

Dr. Madewell and Dr. Charniga are fellows in the Public Health Analytics and Modeling Track of the CDC Steven M. Teustch Prevention Effectiveness Fellowship.

## Acknowledgments

Monkeypox response teams from state and local health departments in the following jurisdictions: California, Chicago, Colorado, District of Columbia, Hawaii, Florida, Illinois, Michigan, New York City, North Carolina, Rhode Island, South Carolina, and Tennessee. Hawaii contributed data, but the case pairs did not meet the inclusion criteria. Center for Forecasting and Outbreak Analytics for technical assistance, Monkeypox States, Tribal, Local and Territorial Support Task Force for outreach and liaison, Data Analytics and Visualization Task Force Informatics Team for data management and support.

## Disclaimer

The findings and conclusions in this report are those of the author(s) and do not necessarily represent the views of the Centers for Disease Control and Prevention.

## Ethics statement

This activity was reviewed by CDC and was conducted consistent with applicable federal law and CDC policy (45 C.F.R. part 46, 21 C.F.R. part 56; 42 U.S.C. Sect. 241(d); 5 U.S.C. Sect. 552a; 44 U.S.C. Sect. 3501 et seq).

## 2022 Monkeypox Outbreak Response Team

CDC: Julia Shaffner

California Department of Public Health: Shua J. Chai, Marisa A.P. Donnelly, Robert E. Snyder, Cameron Stainken, Eric C. Tang, Akiko Kimura, Jason Robert C. Singson, Philip Peters Colorado Department of Public Health & Environment: Robyn Weber and Erin Youngkin District of Columbia Department of Health: Sarah Gillani, Karla Miletti, Allison Morrow Tennessee Department of Health: Caleb Wiedeman

Florida Department of Health: Katharine Saunders, Danielle Stanek, Joshua Moore

Chicago Department of Public Health: Bridget Brassil, Isaac Ghinai, Janna Kerins, Aaron Krusniak, Sarah Love, Peter Ruestow, Emma Weber

North Carolina Department of Health and Human Services: Erin Ricketts

Will CHD Monkeypox Investigation Team

Kendall CHD Monkeypox Investigation Team

## References

1. Centers for Disease Control and Prevention. Monkeypox. 2022 [cited 2022 September 19]; Available from: https://www.cdc.gov/poxvirus/monkeypox/

2. Delaney KP, Sanchez T, Hannah M, Edwards OW, Carpino T, Agnew-Brune C, et al. Strategies Adopted by Gay, Bisexual, and Other Men Who Have Sex with Men to Prevent Monkeypox virus Transmission - United States, August 2022. MMWR Morb Mortal Wkly Rep. 2022 Sep 2;71(35).

3. Reynolds MG, Yorita KL, Kuehnert MJ, Davidson WB, Huhn GD, Holman RC, et al. Clinical manifestations of human monkeypox influenced by route of infection. The Journal of infectious diseases. 2006;194(6):773–80.

4. UK Health Security Agency. Investigation into monkeypox outbreak in England: technical briefing 1. 2022 [cited 2022 August 25]; Available from: https://www.gov.uk/government/publications/monkeypox-outbreak-technical-briefings/investigation-into-monkeypox-outbreak-in-england-technical-briefing-1#:~:text=Without%20correcting%20for%20right%2Dtruncation,8.9)%2C%20see%20Figure%202a.

5. Guzzetta G, Mammone A, Ferraro F, Caraglia A, Rapiti A, Marziano V, et al. Early Estimates of Monkeypox Incubation Period, Generation Time, and Reproduction Number, Italy, May-June 2022. Emerg Infect Dis. 2022 Aug 22;28(10).

6. Gostic KM, McGough L, Baskerville EB, Abbott S, Joshi K, Tedijanto C, et al. Practical considerations for measuring the effective reproductive number, Rt. PLoS Computational Biology. 2020;16(12):e1008409.

7. Cori A, Cauchemez S, Ferguson NM, Fraser C, Dahlqwist E, Demarsh PA, et al. Package ‘EpiEstim’. CRAN: Vienna Austria. 2020.

8. Charniga K, Masters NB, Slayton RB, Gosdin L, Minhaj FS, Philpott D, et al. Estimating the incubation period of monkeypox virus during the 2022 multi-national outbreak. medRxiv. 2022.

9. Du Z, Xu X, Wu Y, Wang L, Cowling BJ, Meyers LA. Serial interval of COVID-19 among publicly reported confirmed cases. Emerging infectious diseases. 2020;26(6):1341.

