## Appendix for "Serial interval and incubation period estimates of monkeypox virus infection in 12 U.S. jurisdictions, May – August 2022"

Supplementary tables

**Table S1.** Type of contact among case pairs included in the serial interval analysis. Demographic information was unavailable for all case pairs. Case pairs with more invasive exposures (i.e., sexual or intimate contact) likely had also additional types of contact.

|  | Symptom onset, N (%) | Rash onset, N (%) |
| --- | --- | --- |
| Caregiving | 1 (1.8) | 0 (0) |
| Face-to-face contact, not including intimate contact | 1 (1.8) | 0 (0) |
| Healthcare | 1 (1.8) | 1 (2.5) |
| Household | 2 (3.5) | 2 (5.0) |
| Shared bedding | 3 (5.3) | 1 (2.5) |
| Sexual or intimate contact | 49 (86.0) | 36 (90.0) |
| Total | 57 (100) | 40 (100) |

**Table S2.** Distributions of estimated incubation period and serial interval of monkeypox virus, United States, May – August 2022. The distributions for the incubation period were log-normal, while the distributions for the serial interval were gamma.

|  | Incubation period | | Serial interval | |
| --- | --- | --- | --- | --- |
|  | Log mean (95% CrI) | Log standard deviation (95% CrI) | Shape (95% CrI) | Scale (95% CrI) |
| Symptom onset | 1.5 (1.2 – 1.8) | 0.7 (0.5 – 1.0) | 2.9 (2.0 – 4.1) | 2.9 (2.0 – 4.4) |
| Rash onset | 1.8 (1.6 – 2.1) | 0.6 (0.4 – 0.8) | 2.8 (1.8 – 4.2) | 2.5 (1.6 – 4.0) |

CrI: credible interval

Supplementary figures

**Figure S1.** Temporal trends in monkeypox case pairs data, United States, May – August 2022.

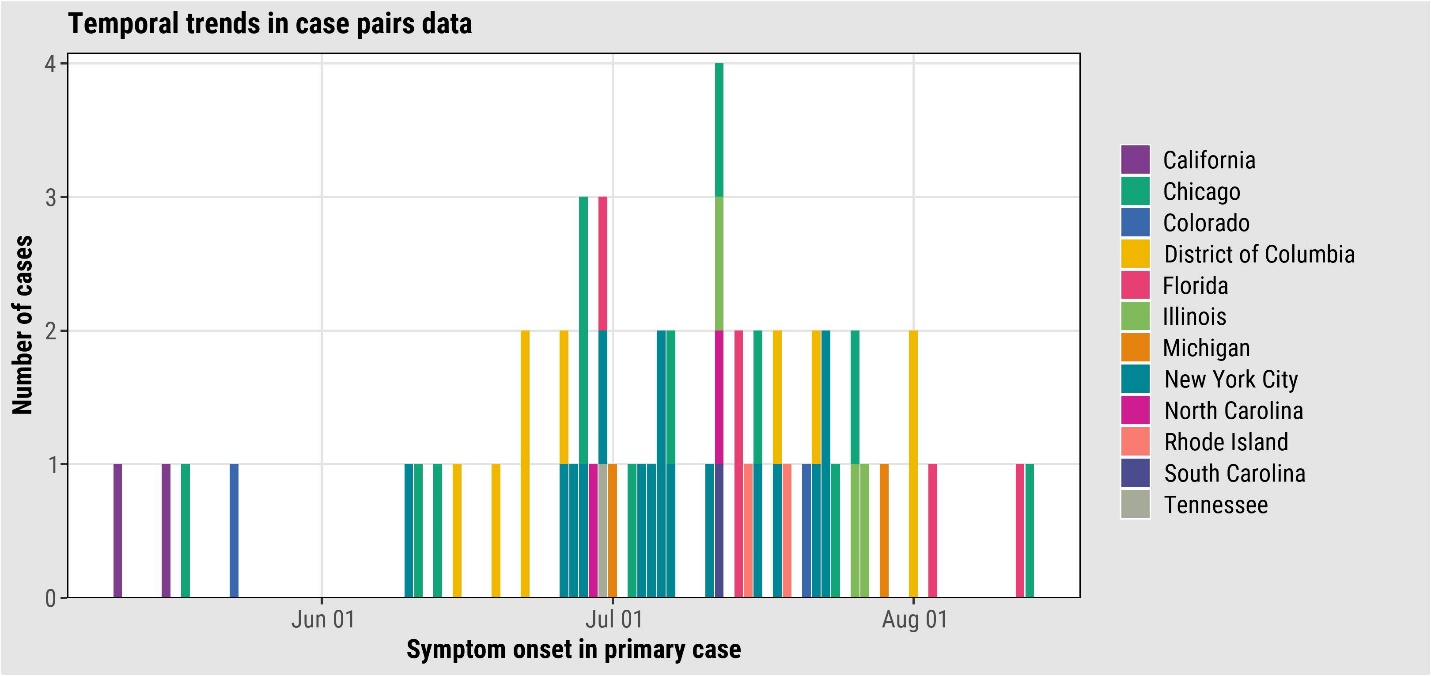

**Figure S2.** Correlation between the serial interval (SI) and incubation period in secondary cases that were included in both analyses (N = 15), United States, May – August 2022. Green points represent the minimum time from exposure to (A) symptom onset or (B) rash onset, blue points represent the maximum time, and purple points represent the midpoint between the minimum and maximum times. Pearson’s correlation coefficient is presented for the purple points. The black reference line is y=x.

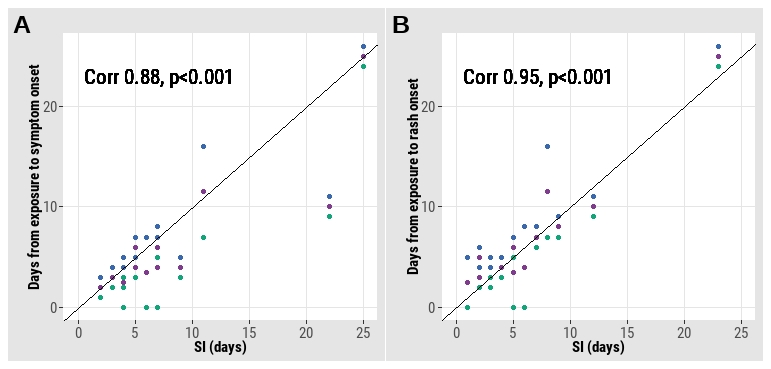
